# Multiplex qPCR Assay for HIV-1 Proviral DNA Detection and Subtype Characterization: Exploiting Quenching of Multiple Fluorophores with a Single Quencher Operating in *Trans*

**DOI:** 10.1101/2025.03.13.25323928

**Authors:** Daisy Patel, Krishna H. Goyani, Isha Sharma, Shalin Vaniawala, Pratap N. Mukhopadhyaya

## Abstract

Accurate detection and quantification of HIV-1 proviral DNA are critical for effective patient monitoring and therapeutic decision-making. In this study, we developed a multiplexed quantitative PCR (qPCR) assay designed to detect HIV-1 proviral DNA, determine viral subtype, specifically identifying the predominant subtype C and validate assay performance using an internal control. Gene-specific primers were engineered by appending an 8-base biotag followed by a common 18-base sequence at the 5′ end, enabling the simultaneous amplification of multiple target sequences. Fluorescent probes labeled with FAM, SUN/VIC, and Cy5 were employed for detection, and a novel strategy involving quenching of labeled probes in trans was implemented to enhance assay flexibility and cost-effectiveness compared to conventional cis-quenched probes. The assay was initially optimized using synthetic linear double-stranded DNA templates representing the HIV-1 gag region, while externally added human chromosomal DNA served as a control for PCR inhibition. Validation was performed on a panel of 11 clinical samples previously analyzed for drug resistance mutations. Results indicated robust amplification of HIV-1 proviral DNA, accurate subtype determination, and reliable internal control performance, with profiles closely matching those obtained by gold standard sequencing-based assays. One sample exhibited PCR inhibition, underscoring the need for internal control monitoring. Overall, the multiplexed qPCR assay provides a sensitive, specific, and efficient tool for comprehensive HIV-1 reservoir quantification and molecular epidemiological studies, potentially informing improved clinical management and personalized treatment strategies. Furthermore, this novel methodology significantly reduces reagent costs and processing time while maintaining high sensitivity, making it ideal for routine clinical and research applications.

## Introduction

The polymerase chain reaction (PCR) technique has proven to be a powerful tool for detecting HIV proviral DNA, offering significant advantages in HIV diagnostics. PCR can detect HIV proviral DNA in various sample types, including peripheral blood mononuclear cells (PBMCs), dried blood spots (DBS), and even brain tissue, with high sensitivity and specificity (Aoki et al., 1990; Bo □Ni et al., 1993; Cassol et al., 1991; Cassol et al., 1992). One of the key advantages of PCR-based HIV proviral DNA detection is its ability to quantify viral load, which can be correlated with disease progression and treatment efficacy. For instance, a study found a significant difference in HIV proviral DNA levels between symptomatic and asymptomatic patients, suggesting that higher viral replication is associated with the onset of clinical symptoms (Genesca et al., 1990). Additionally, PCR can detect HIV proviral DNA in various cell types, including CD4 cells, macrophages, and even sperm cells, providing insights into viral reservoirs and transmission mechanisms (Bagasra et al., 1994; Hufert et al., 1989). However, the genetic variability of HIV-1 can pose challenges for PCR-based diagnostics, particularly with African isolates (Candotti et al., 1991). To address this issue and ensure reliable results across different laboratories, standardized commercial assays and quality assurance programs have been developed (Jackson et al., 1993). Furthermore, advancements such as competitive PCR-based assays have improved the quantitative detection of HIV-1 viremia, cellular transcripts, and proviral DNA sequences (Menzo et al., 1992). These developments highlight the ongoing need for refining PCR techniques to enhance HIV diagnostics and monitor treatment effectiveness.

HIV-1 subtype C viruses are associated with nearly half of worldwide HIV-1 infections and are most predominant in India and parts of Africa (Siddappa et al., 2004). In India, studies have shown that subtype C is the most prevalent, with 78.4% of samples analyzed being of this subtype (Sahni et al., 2002). This prevalence has significant implications for therapeutic decisions and vaccine development in the region. Interestingly, all patients in a study of antiretroviral drug-naive individuals in southern India had at least one protease and/or RT mutation at a known subtype B drug resistance position (Balakrishnan et al., 2005). This highlights the need for careful monitoring of drug resistance, even in treatment-naive individuals. Furthermore, a case report from southern Africa, where HIV-1 subtype C also predominates, demonstrated the development of multidrug resistance mutations to four antiretroviral classes in a patient on dolutegravir-based therapy (Seatla et al., 2018). In conclusion, the high prevalence of HIV-1 subtype C in India necessitates specific diagnostic and therapeutic strategies. The development of subtype C-specific PCR methods, such as C-PCR, can facilitate rapid molecular epidemiologic characterization (Siddappa et al., 2004). Additionally, the identification of conserved signature amino acid positions in Indian subtype C viruses could assist in epidemiologic tracking and vaccine development (Shankarappa et al., 2001). Given the potential for drug resistance, even in treatment-naive individuals, there is a clear need for ongoing surveillance and resistance testing to guide therapeutic decisions in regions where subtype C predominates (Hirsch et al., 2008).

In this study, a lesser-known technique of single-tube multiplexing of individual target amplification on a real-time PCR platform was employed. This approach offered advantages in terms of cost-effectiveness and license-free use of dye-labeled probes while demonstrating assay performance similar to that achieved by dual-labeled TaqMan probes. Additionally, contact quenching was incorporated within this framework to enhance fluorescence signal detection and minimize background noise. HIV-1 proviral DNA was selected as the analyte of choice due to its ease of extraction from clinical samples and the availability of internal control sets embedded within the DNA. These internal controls served as PCR inhibition control features, ensuring the reliability and accuracy of the assay.

## Material and methods

### Research Design

This study employed a combination of end-point PCR and real-time PCR techniques to optimize and detect HIV proviral DNA, specifically targeting the gag gene and C subtype-specific region.

### Research Method

The methodology involved optimizing each target for annealing temperature and primer concentration individually before multiplexing. End-point PCR products were detected using gel electrophoresis, while qPCR utilized fluorescent multiplexing for real-time detection.

### Literature Review

A comprehensive literature review was conducted to identify existing methodologies and best practices for detecting HIV proviral DNA using PCR techniques.

### Study Samples

Synthetic DNA targets were used for the initial optimization of the assay to ensure accurate amplification and detection. For validation, archived HIV proviral DNA samples (n=11) were obtained from an NABL-accredited laboratory. These archived samples provided a reliable reference for evaluating the assay’s performance under clinically relevant conditions.

### Authentication of Human DNA Extracted from Peripheral Blood

Human peripheral blood DNA was authenticated by amplifying a 437 bp region of the GAPDH gene using forward (5′-TGAAGGTCGGAGTCAACGGATTTGGT-3′) and reverse (5′-CATGTGGGCCATGAGGTCCACCAC-3′) primers, with 0.4 µM of each primer, 1X PCR master mix, and 75 ng of extracted DNA under thermal cycling conditions of 94°C for 2 min, followed by 35 cycles of 94°C for 30 s, 60°C for 30 s, 72°C for 1 min, and a final extension at 72°C for 2 min; amplicons were resolved on a 2% agarose gel and visualized under UV after ethidium bromide staining (0.5 µg/mL).

### Data Collection

Archived DNA provided by a NABL-accredited laboratory for this retrospective study was used, and PCR assays were performed to detect the targeted HIV proviral DNA regions. Data from end-point PCR were collected as images of agarose gel electrophoresis runs, while qPCR data were recorded as fluorescent signals captured by the software during the extension step of each cycle.

### End-Point PCR Optimization

To optimize the end-point PCR conditions for HIV-1 proviral DNA detection, a series of reactions were performed using synthetic linear HIV-1 proviral DNA as the template. The PCR reactions were carried out in two steps. In the first step, a primary PCR reaction was set up using oligonucleotide primers targeting positions 130–157 and 1075–1104 of the HXB2 genome (Siddappa et al., 2004). The reaction mixture consisted of 10 pmol of each primer, 1X PCR master mix (Thermo Fisher Scientific), and 4 µL of template DNA (5-10 ng synthetic linear double-stranded HIV-1 proviral DNA appended with 75 ng of human DNA in 1 µL and extracted from peripheral blood of a healthy individual) in a final volume of 25 µL. The thermal cycling conditions were as follows: Initial denaturation at 95°C for 5 minutes, followed by 35 cycles of 95°C for 30 seconds, 60°C for 30 sec and 72°C for 30 sec. The amplified product from this reaction was not resolved on an agarose gel but was directly used as a template for the second-stage PCR.

In the second-stage PCR, a 25 µL reaction mixture was prepared, consisting of 2 µL of the first-stage PCR product, 1X PCR master mix, and varying concentrations of forward and reverse primers (0.2 µM, 0.4 µM, and 0.8 µM) to determine the optimal primer concentration. The best amplification efficiency was observed with 0.4 µM of each primer, which was subsequently selected for further reactions. The thermal cycling conditions included 35 cycles, each consisting of 94°C for 1 minute, 70°C for 40 seconds, and 72°C for 40 seconds. Furthermore, annealing temperatures of 68°C, 70°C, and 72°C were tested to optimize amplification conditions. The optimal annealing temperature was determined to be 70°C, as it yielded the strongest and most specific amplification. This optimized protocol was used for all subsequent end-point PCR reactions.

### Design of gene-specific primers & contact quenching probes

The gene-specific primers were designed by engineering a biotag sequence followed by a dye-labeled probe-hybridizing stretch attached to the 5′ end of the gene-specific primers. All primers were designed based on the HXB2 HIV-1 genome sequence coordinates. In Stage 1 PCR, primers targeting positions 130 to 157 (forward primer) and 1075 to 1104 (reverse primer) (Siddappa et al., 2004) were used. These primers did not include any engineered regions at their 5′ ends.

In Stage 2 nested PCR, the primers were as follows (5’ – 3’):

- HIV-1 gag gene-specific primers:
  - o Forward primer: 683 to 709; Reverse primer: 890 to 915
- Subtype C-specific primers:
  - Forward primer: 256 to 284; Reverse primer: 367 to 393
- GAPDH gene primers
  - GAPDH F: CCCCACACACATGCACTTAC; GAPDH R: TCCCTCCTACAAAAGGGACAGAG

The gene-specific primers were thereafter engineered by appending a common 18 □bp sequence (“GCGGACTGGGTAAACGTC”) upstream of an 8□bp biotag, with the biotag sequences being “CGTACGGC” for the HIV-1 gag gene-specific forward primer, “GGACGGTG” for the subtype C-specific forward primer, and “GGACAACG” for the GAPDH gene primer; furthermore, the fluorescent probes for FAM, SUN (VIC), and Cy5 were designed to be homologous to this 18□bp sequence—with the FAM probe incorporating an additional 8 bp segment as mentioned above and matching the gag biotag at its 3′ end, and the SUN (VIC) and Cy5-labeled probes containing biotags corresponding to the subtype C-specific region and human GAPDH gene, respectively, at their 3′ end. All dye-labeled probes were quenched in trans using an 18□bp oligonucleotide fully complementary to the common 18□bp sequence and tagged with Black Hole Quencher II at its 3′ end.

### qPCR Optimization

For qPCR optimization, the reaction conditions determined to be optimal during end-point PCR were used, with modifications specific to the qPCR workflow. The PCR master mix used for qPCR was a commercially available formulation procured from Thermo Fisher, USA, and was used at a final concentration of 1X in a 25 µL reaction volume. However, in all cases, first-stage end-point PCR was also performed, and 2 µL of its product was used for qPCR reactions. Briefly, 2 µL of the first-stage PCR product was added to a PCR master mix to achieve a final concentration of 1X. Each oligonucleotide primer (three sets of primer pairs—for the *gag* gene, the HIV-1 subtype C-specific region, and the human *GAPDH* gene) was added at a final concentration of 0.4 µM. Additionally, 0.2 µM of each dye-labeled probe—FAM, SUN (VIC), and Cy5—was included, along with a single common quencher-labeled probe (Black Hole Quencher II) at a quantity of 0.2 µM. The final reaction volume was 25 µL. All assays were run on a QuantStudio 5 real-time PCR machine. Similar to end-point PCR, each target was individually optimized before multiplexed detection. Synthetic HIV-1 proviral linear DNA was used for optimization. The finalized optimized conditions were then applied to analyze a panel of 11 archived HIV-1 proviral DNA samples, which had been previously tested for HIV-1 proviral DNA positivity and subtyping (C or non-C). Additionally, the samples were also assessed for PCR inhibition. The thermal cycling conditions consisted of an initial hold at 65°C for 15 minutes and 95°C for 2 minutes, followed by 10 cycles of 95°C for 10 seconds, 64°C for 20 seconds, and 68°C for 20 seconds. During these initial 10 cycles, no fluorescence data was collected. This was followed by 25 cycles, each consisting of 95°C for 10 seconds, 57.1°C for 30 seconds, and 68°C for 30 seconds. In these 25 cycles, fluorescence data was collected during the extension step of each cycle. Cumulative signals in the FAM, SUN, and Cy5 channels were analyzed by studying the graphs generated by the qPCR software, and inferences were accordingly drawn based on the observed data. The threshold was set to automatic mode in the QuantStudio 5 qPCR machine. Graphs crossing the threshold before the end of the cycling process were considered positive, while those that remained below the threshold were considered negative.

### Ethics Statement

This study was approved by the Wobble Base Bioresearch Ethics Committee (Approval No. WBBPL/EC/Mar2005/001) and conducted in accordance with ethical guidelines.

### Age Statement

Participant ages are reported in ranges of at least five years to maintain anonymity.

## Results

Human DNA was extracted from peripheral blood to serve as an additional template alongside the synthetic linear HIV proviral DNA fragment, mimicking clinical human proviral DNA. The DNA quantity was confirmed by PCR amplification of a 437 bp region of the human GAPDH gene, successfully detected, validating the authenticity of the extracted DNA (Figure 1).

**Figure 1:**
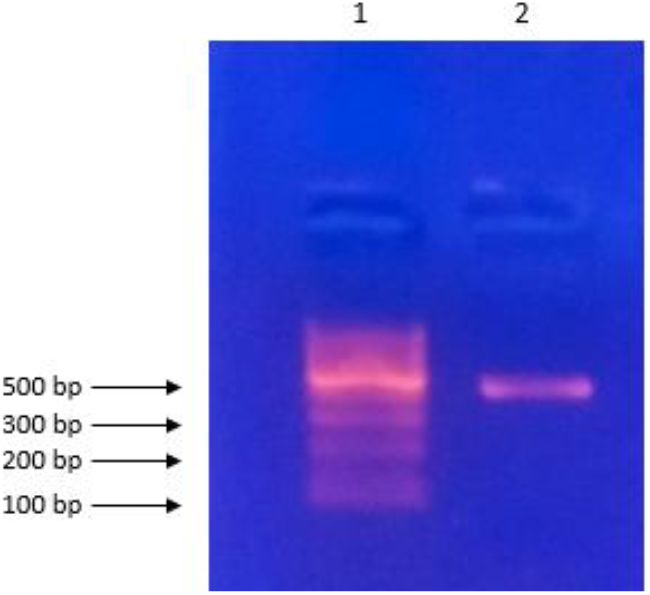
Agarose gel electrophoresis showing successful amplification of a 437 bp PCR amplicon generated from the human GAPDH gene when DNA extracted from human peripheral blood was used as a template. Lane 1: 100 bp DNA size standard; Lane 2: Human peripheral blood DNA amplified with human GAPDH-specific primers to generate the desired 437 bp PCR amplicon.

Optimization of the thermal amplification of the three individual targets—namely, the HIV gag gene, the C subtype-specific region, and the human GAPDH gene—was evident from the singular bands detected on the agarose gel (Figure 2a, 2b, and 2c, respectively). Post multiplexing of all three targets, the data indicate that the concentration of all three sets of primers plays a pivotal role in generating bands for each target. Primer concentrations of 5 and 20 pmol in a 20 µL PCR reaction volume was found to be suboptimal, whereas a concentration of 10 pmol for all three sets of primers (each pair targeting the HIV gag gene, C subtype-specific region, and human GAPDH gene) generated the best amplification profile (Figure 2d). This established the optimal concentration of the non-labeled primers for the qPCR protocol.

**Figure 2a:**
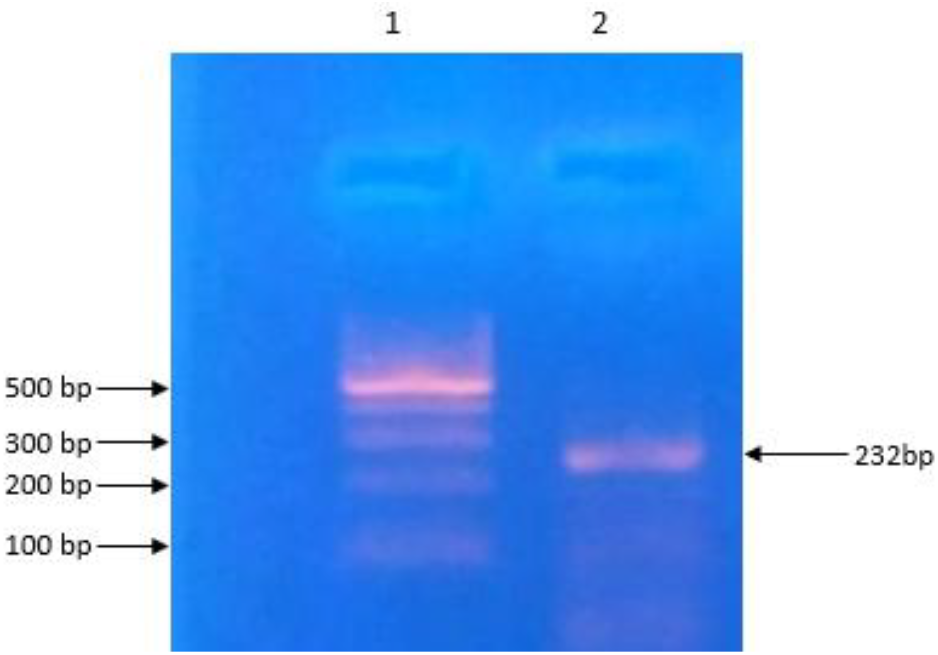
Agarose gel electrophoresis showing optimized amplification of a 232 bp fragment of the HIV-1 gag gene when synthetic linear double-stranded HIV-1 proviral DNA was used as a template.

**Figure 2b:**
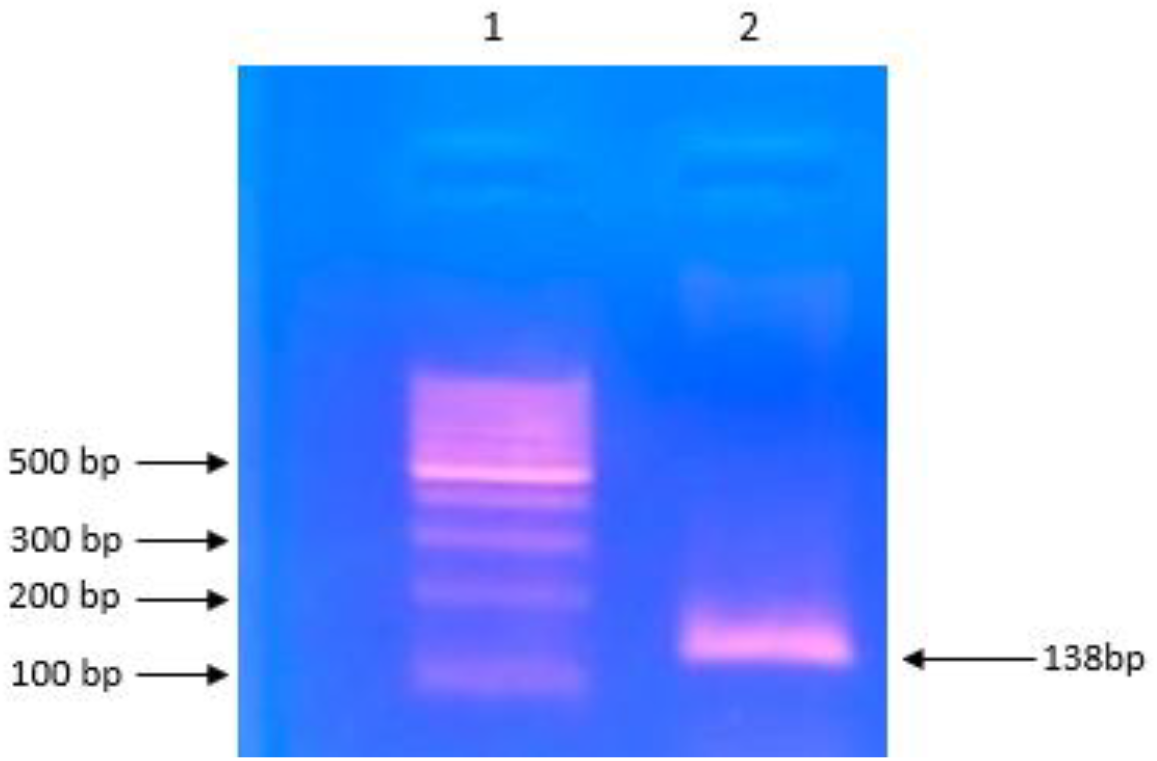
Agarose gel electrophoresis showing optimized amplification of a 138 bp fragment specific to the HIV-1 C subtype variant when synthetic linear double-stranded HIV-1 proviral DNA was used as a template.

**Figure 2c:**
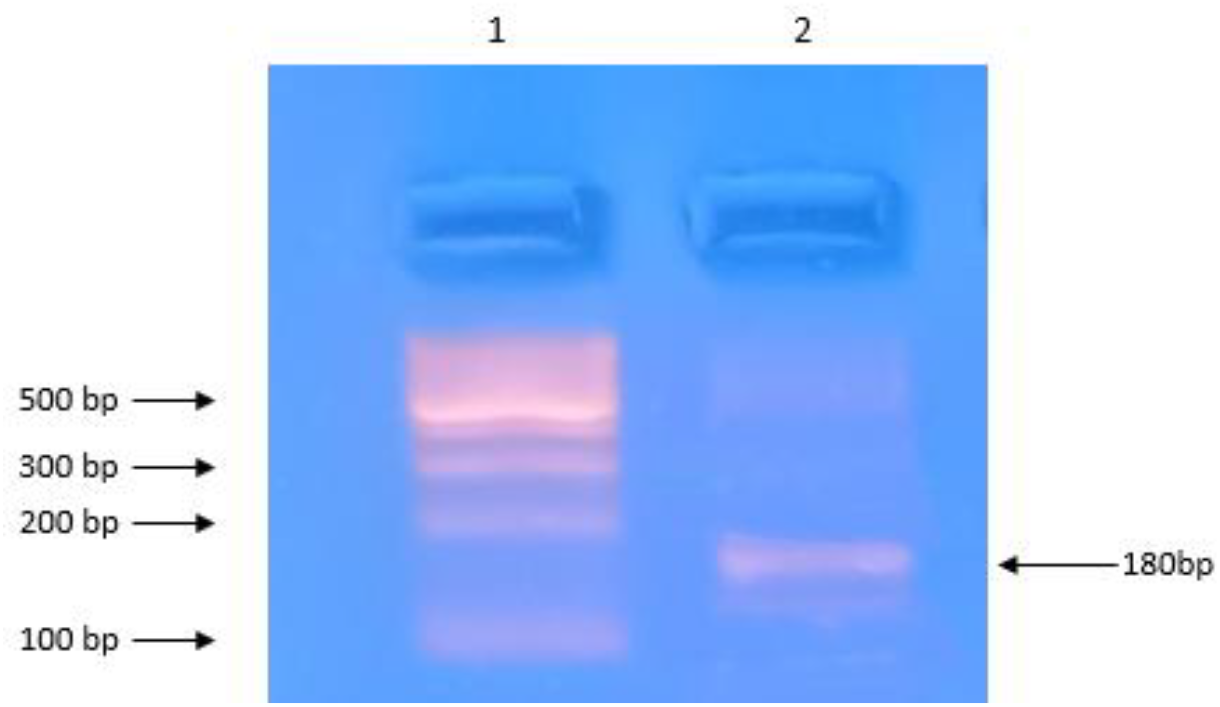
Agarose gel electrophoresis showing optimized amplification of a 180 bp fragment of the human GAPDH gene when synthetic linear double-stranded HIV-1 proviral DNA, along with externally added human peripheral blood DNA, was used together as a template.

**Figure 2d:**
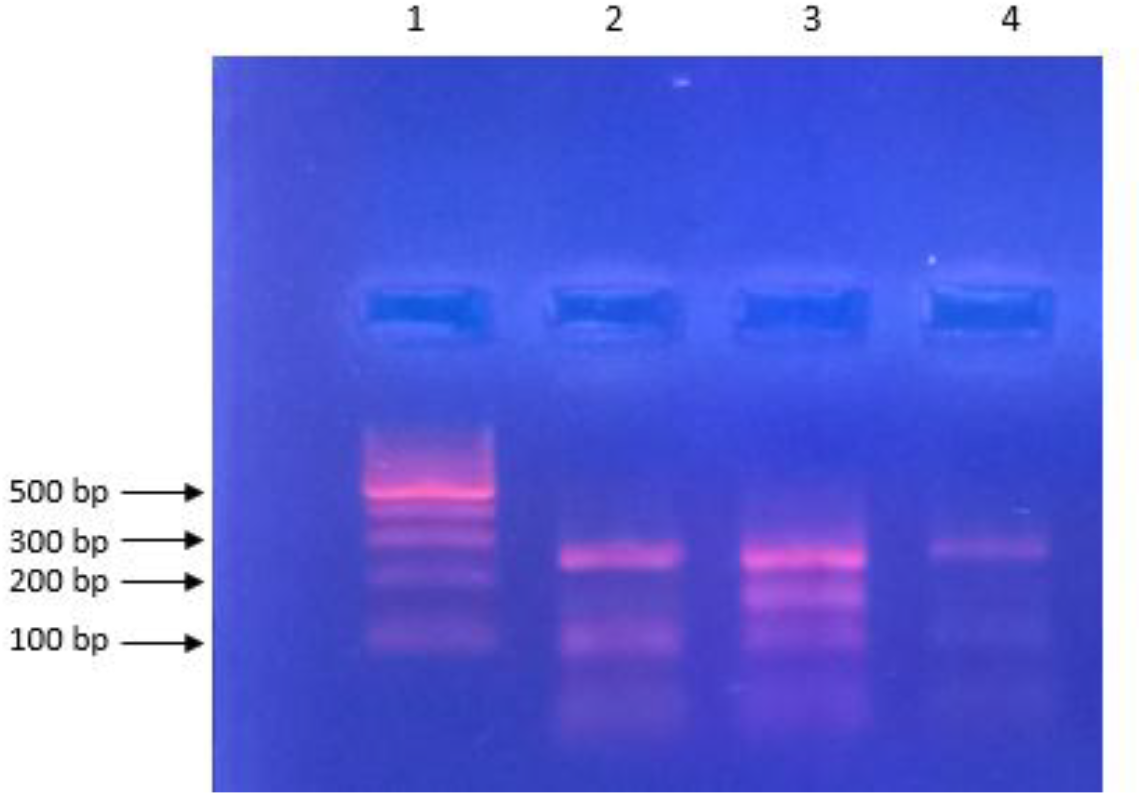
Agarose gel electrophoresis of the multiplex endpoint PCR reaction showing PCR efficiency at different primer concentrations for each of the three targets in a 25 µL reaction volume: (a) 5 pmoles, (b) 10 pmoles, and (c) 20 pmoles. Lane 1: 100 bp DNA size standard; Lane 2: PCR with 5 pmoles of primer concentration; Lane 3: PCR with 10 pmoles of primer concentration; Lane 4: PCR with 20 pmoles of primer concentration. The three amplicons visible in the reaction with 10 pmoles of primer concentration for each of the three primer pairs (Lane 3) correspond to the HIV-1 gag gene (258 bp), the HIV-1 C subtype-specific region (164 bp), and a region within the human GAPDH gene (187 bp).

The qPCR data from different template combinations yielded the desired results in both monoplex and multiplex reactions. Table 1 summarizes the various template combinations used in this study along with their corresponding fluorescence signal profiles obtained during qPCR optimization. Figure 3 (a–d) graphically represents the different templates tested during the optimization process. The curve profiles observed in these figures are explained in the corresponding legends of each figure.

**Table 1:**
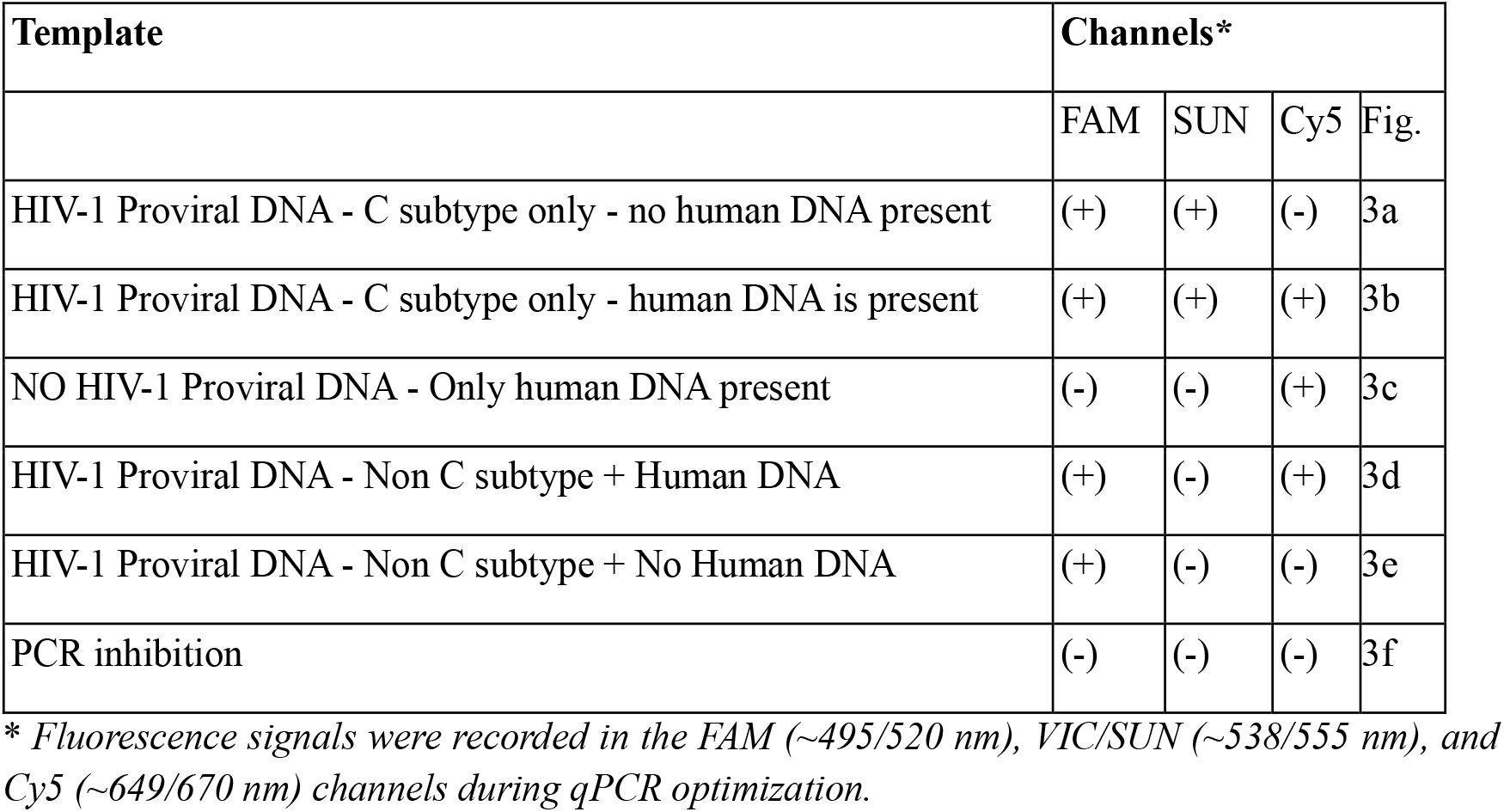
Various template combinations and corresponding fluorescence recorded in qPCR during the optimization protocol for multiplex detection of HIV-1 and its C subtype variant. In all cases, the HIV-1 proviral DNA template was represented by a synthetically generated linear DNA obtained from the diagnostically relevant region of the HIV-1 proviral genome of C and non-C subtype variants.

| Template | Channels* |  |  |  |
| --- | --- | --- | --- | --- |
|  | FAM | SUN | Cy5 | Fig. |
| HIV-1 Proviral DNA - C subtype only - no human DNA present | (+) | (+) | (-) | 3a |
| HIV-1 Proviral DNA - C subtype only - human DNA is present | (+) | (+) | (+) | 3b |
| NO HIV-1 Proviral DNA - Only human DNA present | (-) | (-) | (+) | 3c |
| HIV-1 Proviral DNA - Non C subtype + Human DNA | (+) | (-) | (+) | 3d |
| HIV-1 Proviral DNA - Non C subtype + No Human DNA | (+) | (-) | (-) | 3e |
| PCR inhibition | (-) | (-) | (-) | 3f |

\* Fluorescence signals were recorded in the FAM (~495/520 nm), VIC/SUN (~538/555 nm), and Cy5 (~649/670 nm) channels during qPCR optimization.

**Figure 3a:**
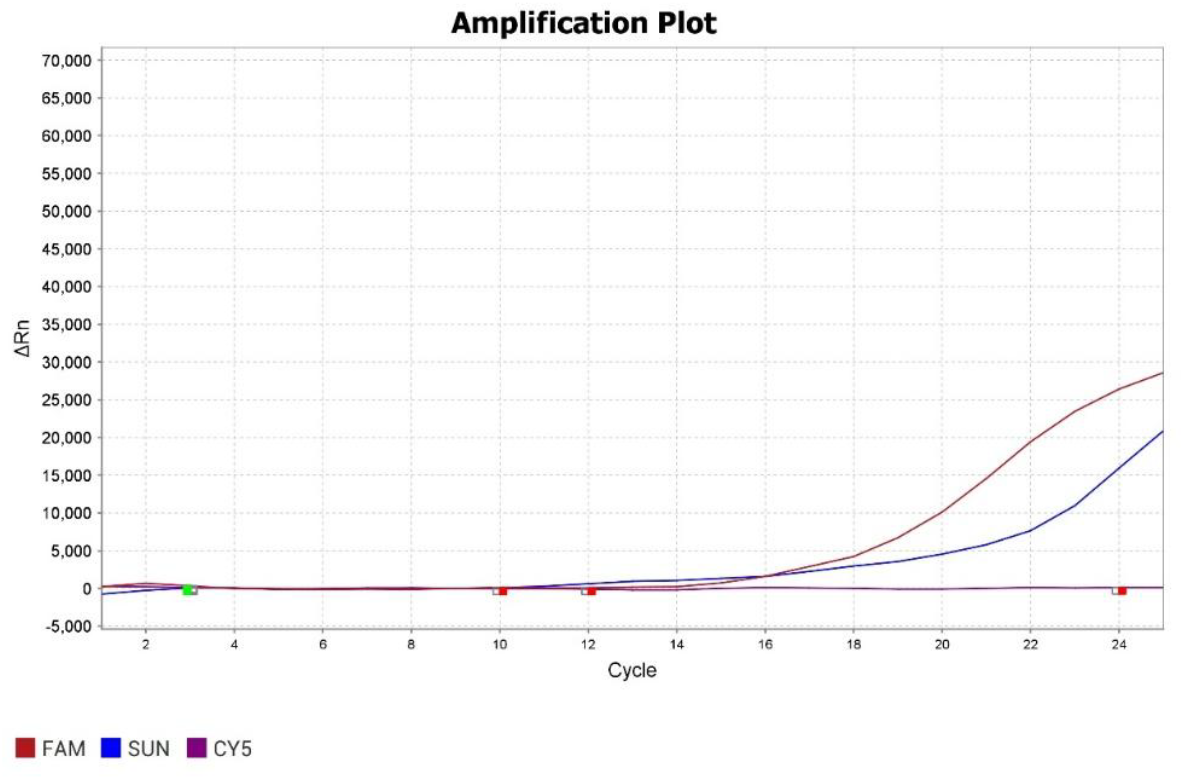
qPCR data from a run where the template comprised a synthetic linear double-stranded DNA representing the gag region of an HIV-1 C subtype variant but no externally added human chromosomal DNA. A signal was detected in the FAM (∼495/520 nm) channel, indicating the presence of the HIV-1 gag gene, and in SUN/VIC channel (538/555 nm) indicating that the HIV-1 variant. But no Cy5 (∼649/670 nm) signal was detected since no externally added human DNA was there in the reaction environment.

**Figure 3b:**
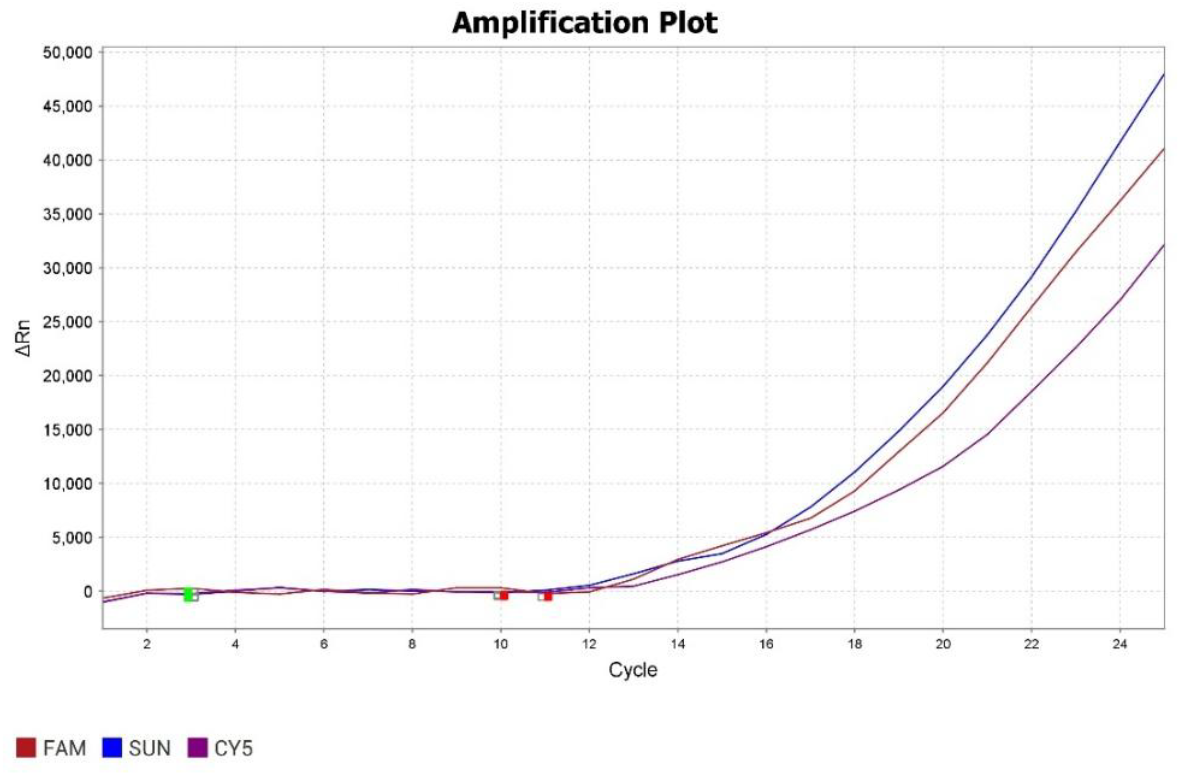
qPCR data from a run where the template comprised synthetic linear double-stranded DNA representing the gag region of an HIV-1 C subtype variant, along with externally added human chromosomal DNA. Signals were detected in the FAM (∼495/520 nm), SUN/VIC (∼538/555 nm), and Cy5 (∼649/670 nm) channels, indicating the presence of the HIV-1 gag gene and the C subtype variant, with no evidence of PCR inhibition.

**Figure 3c:**
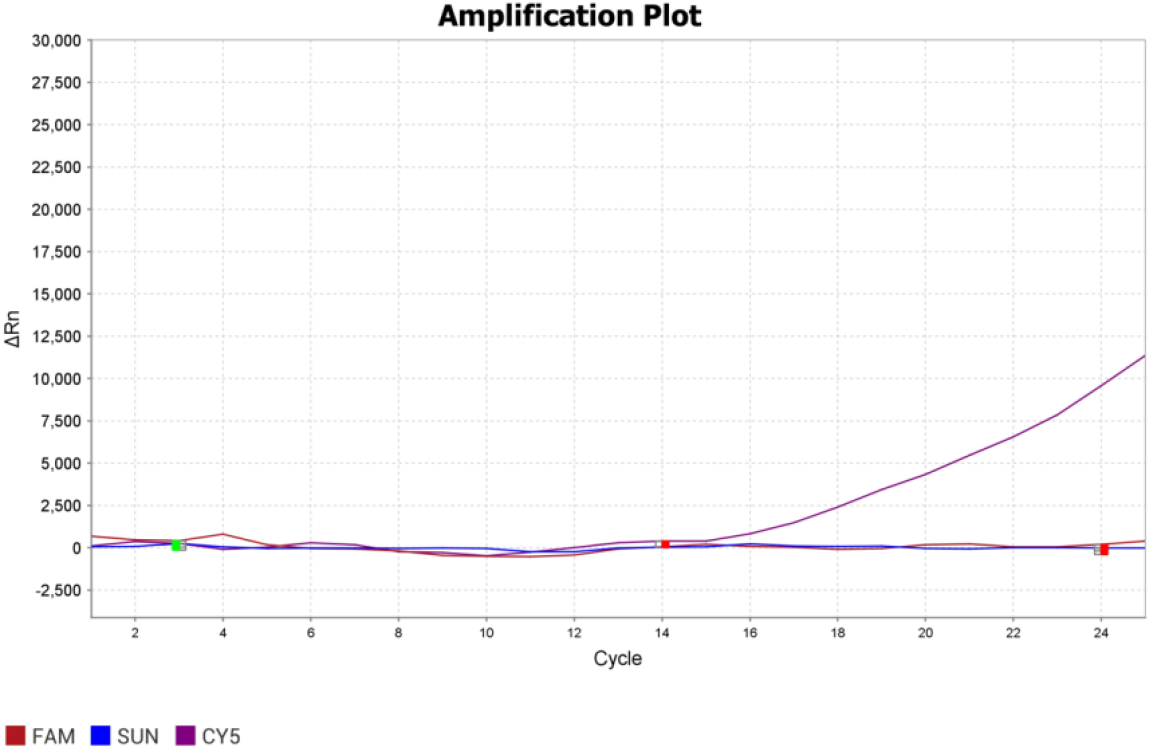
qPCR data from a run where the template comprised only human chromosomal DNA extracted from peripheral blood. A signal was detected only in the Cy5 (∼649/670 nm) channel, confirming that no PCR inhibition occurred during the reaction. This profile corresponds to a clinical sample from a healthy, non-infected individual.

**Figure 3d:**
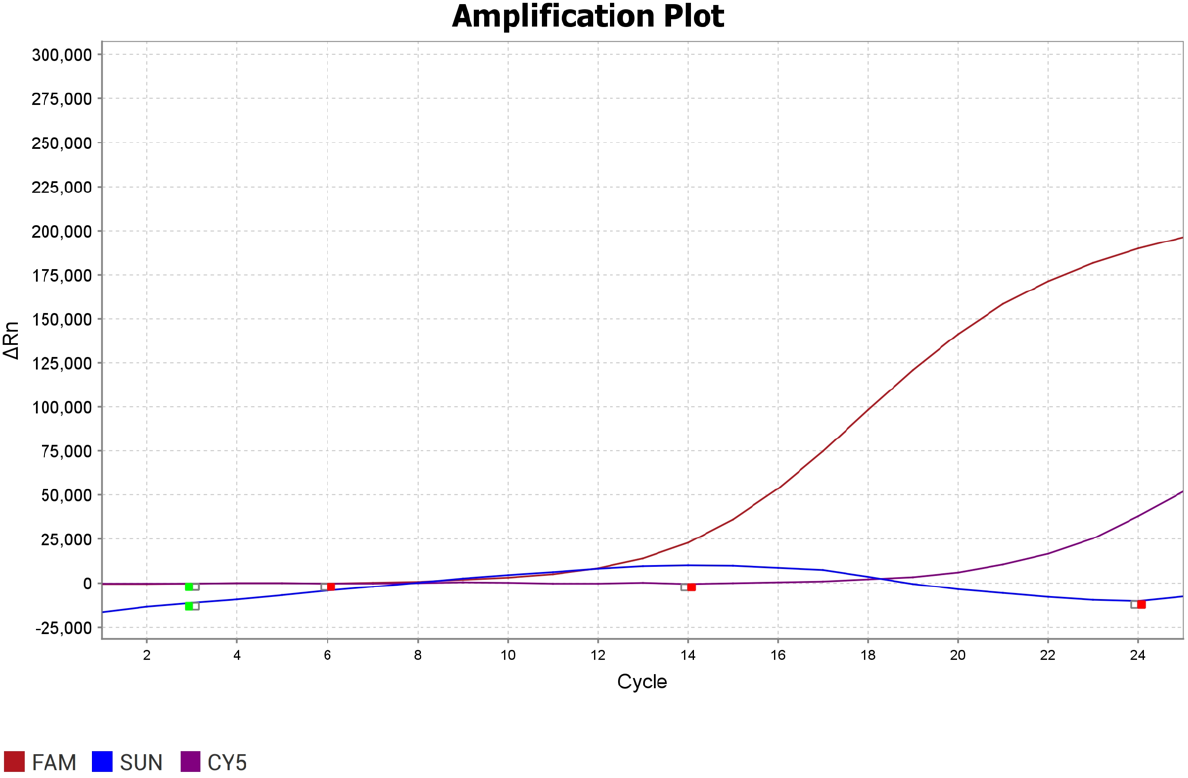
qPCR data from a run where the template comprised a synthetic linear double-stranded DNA representing the gag region of an HIV-1 B subtype variant, along with externally added human chromosomal DNA extracted from peripheral blood. A signal was detected in the FAM ((∼495/520 nm) channel, indicating the presence of the HIV-1 gag gene, and in the Cy5 (∼649/670 nm) channel, confirming that no PCR inhibition occurred during the reaction. No signals were detected for the C subtype-specific region (SUN/VIC channel; 538/555 nm) indicating that the HIV-1 variant represented by the synthetic DNA originated from a non-C (B) subtype of HIV-1 clinical isolate.

**Figure 3e:**
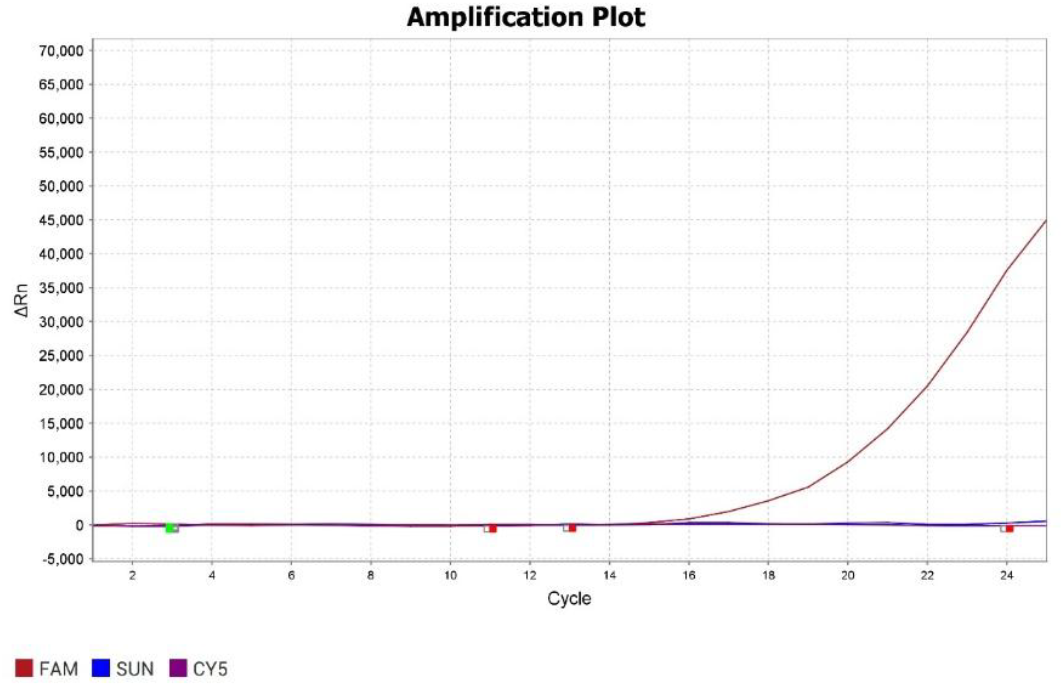
qPCR data from a run where the template comprised synthetic linear double-stranded DNA representing the gag region of an HIV-1 non-C (B) subtype variant, but with no externally added human chromosomal DNA. Signals were detected in the FAM (∼495/520 nm) but no signal was detected in the SUN/VIC (∼538/555 nm), and Cy5 (∼649/670 nm) channels, indicating the presence of the HIV-1 gag gene and absence of C subtype variant specific DNA sequence, with no data to predict PCR inhibition.

**Figure 3f:**
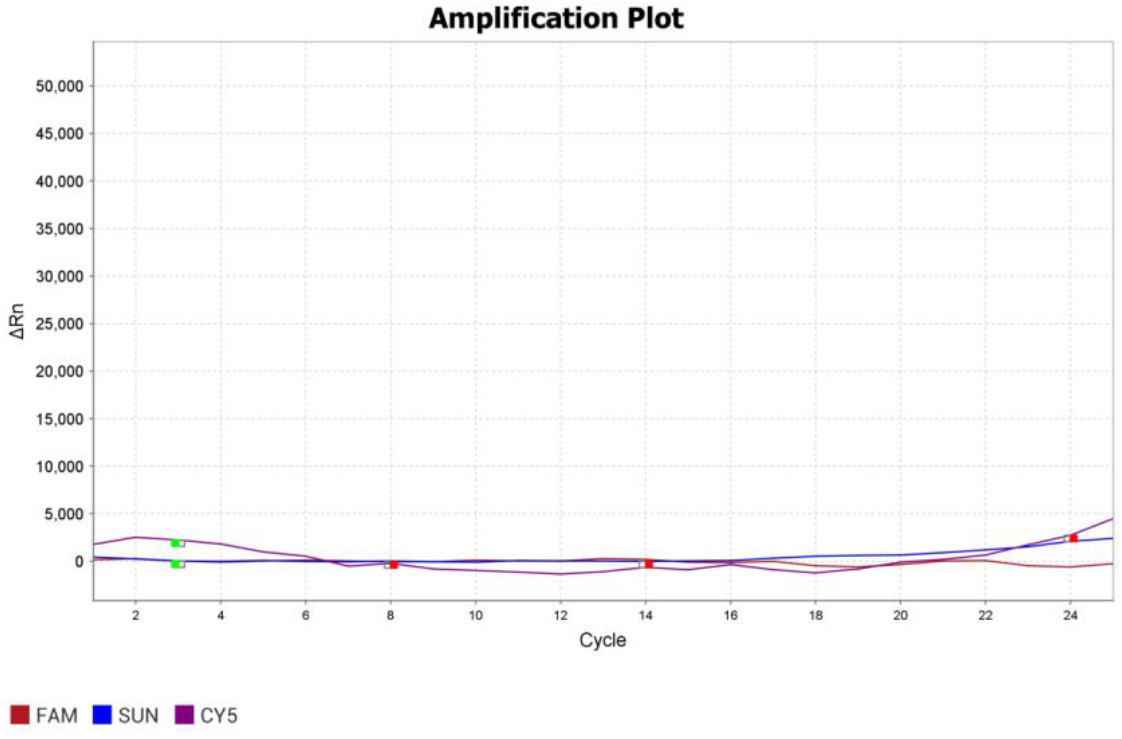
qPCR data from a run where the template comprised a synthetic linear double-stranded DNA representing the gag region of an HIV-1 C subtype variant, along with externally added human chromosomal DNA extracted from peripheral blood containing a significant quantity of PCR inhibitory substances. No signal was detected in the FAM (∼495/520 nm), Cy5 (∼649/670 nm), or SUN/VIC (∼538/555 nm) channels, indicating that despite the presence of an amplifiable quantity of template DNA, the thermal cycling reaction could not take place due to interference from PCR inhibitory substances present in the reaction solution.

The optimized protocol developed in this study was used to validate a set of 11 archived samples obtained from a NABL-accredited laboratory, which were previously tested for HIV-1 proviral DNA positivity. As described above, 2 µL of extracted proviral DNA was used to set up the first-stage PCR reaction on an end-point PCR platform. Subsequently, 5 µL of this first-stage PCR product was used for the qPCR reaction, which comprised three sets of primer pairs corresponding to three different targets, as mentioned in the Materials and Methods section. The qPCR graphs generated for positive reactions were carefully analyzed, and data were recorded based on the Ct values detected for each fluorescence channel. The results are summarized in Table 2 & Supplementary data 1.

**Table 2:** Results obtained from a validation study on a panel of 11 HIV-1 proviral DNA samples previously tested using a published HIV-1 drug resistance analysis protocol for subtype determination. The signal profiles indicate the fluorescence data recorded using a QuantStudio 5 qPCR platform. Raw data pertaining to this summary are available in Supplementary Information 1.

| Sample No. | Signal profile | Confirmed genotype* |
| --- | --- | --- |
| 1 | FAM (+); SUN (+); Cy5 (+) | HIV-1; C Subtype |
| 2 | FAM (+); SUN (-); Cy5 (+) | HIV-1; B Subtype |
| 3 | FAM (+); SUN (+); Cy5 (+) | HIV-1; C Subtype |
| 4 | FAM (+); SUN (+); Cy5 (+) | HIV-1; C Subtype |
| 5 | FAM (+); SUN (+); Cy5 (+) | Healthy Individual |
| 6 | FAM (+); SUN (-); Cy5 (+) | HIV-1; B Subtype |
| 7 | FAM (+); SUN (+); Cy5 (+) | HIV-1; C Subtype |
| 8 | FAM (-); SUN (-); Cy5 (-) | HIV-1; C Subtype |
| 9 | FAM (+); SUN (-); Cy5 (+) | HIV-1; B Subtype |
| 10 | FAM (+); SUN (+); Cy5 (+) | HIV-1; C Subtype |
| 11 | FAM (+); SUN (+); Cy5 (+) | HIV-1; C Subtype |

\* Protocol followed - Acharya et al., 2014

## Discussion

HIV incidence in India has shown significant variations across different populations and time periods: In the early 1990s, HIV incidence was estimated to be rapidly increasing, with the number of infections rising from a few thousand to millions within a decade (Sood et al., 2006). A study in Pune found an HIV incidence rate of 18.6% per year among patients attending sexually transmitted disease clinics, suggesting rapid growth of the epidemic (Brookmeyer et al., 1995). However, more recent data indicate a declining trend in HIV incidence nationally. The age-standardized incidence rate decreased from its peak in 1997 (38.0 and 27.6 per 100,000 for males and females respectively) to 5.4 and 4.6 per 100,000 in 2019 (Shri et al., 2023). Interestingly, while overall incidence has declined, there are significant disparities across populations and regions. A study of men who have sex with men (MSM) in 12 Indian cities found an annualized HIV incidence of 0.87%, ranging from 0-2.2% across sites (Solomon et al., 2015). Among people who inject drugs (PWIDs), a median incidence of 2.9 per 100 person-years was observed across 15 cities, with higher incidence in north/central sites compared to northeastern states (Lucas et al., 2015).

While India has made progress in reducing overall HIV incidence, certain high-risk populations continue to experience elevated rates of new infections. Targeted interventions focusing on key populations like MSM, PWIDs, and female sex workers, along with their partners, remain crucial for further reducing HIV transmission in India (Vickerman et al., 2010). Continued monitoring of incidence patterns across different groups and regions is essential for guiding prevention efforts.

Proviral DNA detection using end-point PCR has been widely employed for diagnosing HIV-1 infection. This method involves amplifying specific regions of the HIV-1 genome integrated into host cell DNA (Jangam et al., 2009; Menzo et al., 1992). The technique has shown high sensitivity and specificity, with the ability to detect as few as 20 copies of HIV-1 proviral DNA from 100 μl of blood (Jangam et al., 2009). Interestingly, PCR-based detection of HIV-1 proviral DNA has revealed genetic differences between viral populations in different biological compartments, such as blood and cerebrospinal fluid, suggesting the existence of specific neurotropic HIV variants (Steuler et al., 1992). Additionally, quantitative PCR assays have demonstrated a correlation between HIV-1 proviral DNA levels and disease progression, with higher levels observed in symptomatic patients compared to asymptomatic individuals (Berry et al., 1994; Genesca et al., 1990). In summary, end-point PCR for HIV-1 proviral DNA detection has proven to be a valuable tool in HIV diagnostics, particularly for infant diagnosis in resource-limited settings (Jangam et al., 2009). The method’s high sensitivity and specificity, combined with its ability to provide quantitative information on viral load and genetic diversity, make it an essential technique in HIV research and clinical management.

In this study, the technology developed was primarily aimed at detecting HIV-1 proviral DNA from human peripheral blood. While one key disadvantage of this platform is its inability to identify positive samples during the window period, the major limitation is the economic feasibility of confirming HIV-1 in the human system using peripheral blood as the analyte.

PCR detection of HIV-1 proviral DNA is the recommended method for diagnosing HIV-1 infection in infants born to HIV-positive mothers, especially in resource-limited settings (Jangam et al., 2009). This method demonstrates high sensitivity and specificity, allowing detection of as few as 20 copies of HIV-1 proviral DNA from 100 μl of blood (Jangam et al., 2009). Interestingly, HIV-1 proviral DNA and antibodies are typically only detectable in infants after 18 months of age, suggesting that mother-to-child transmission often occurs postnatally, likely through breastfeeding (Nyambi et al., 1996). The timing of PCR testing is crucial, as early testing within the first few days of life can identify in utero infections, while later testing at 10-15 days postpartum can detect intrapartum transmissions (Cassol et al., 1994).

This study aimed to develop a multiplexed detection system for HIV-1 in its proviral state, coupled with the identification of subtype C and non-C variants. HIV-1 subtype C is the predominant strain in India, accounting for nearly half of worldwide HIV-1 infections (Shankarappa et al., 2001; Siddappa et al., 2004). A PCR-based strategy (C-PCR) has been developed to specifically identify subtype C viruses, facilitating rapid molecular epidemiologic characterization (Siddappa et al., 2004). This method is particularly relevant for India, as it can aid in vaccine and therapeutic strategies tailored to the country’s epidemic. The C(IN) lineage, a distinct phylogenetic group within subtype C, is prevalent in India and likely descended from a single introduction (Shankarappa et al., 2001). This lineage has unique signature amino acid substitutions, which could assist in epidemiologic tracking and vaccine development specific to Indian HIV-1 subtype C (Shankarappa et al., 2001).

HIV-1 subtype C and non-C subtypes exhibit differences in response to antiretroviral therapy. Subtype C shows a lower rate of accumulation of drug resistance mutations compared to subtype B (Soares et al., 2007). For protease inhibitors, 26% of subtype B strains were resistant, versus only 8% in subtype C. For nucleoside reverse transcriptase inhibitors, 54% of subtype B strains were resistant compared to 23% of subtype C (Soares et al., 2007). Interestingly, while subtype C appears to develop fewer drug resistance mutations, it may have equal transmission fitness but reduced pathogenic fitness relative to other group M HIV-1 isolates (Abraha et al., 2009). This could explain its global prevalence despite potentially lower drug resistance. Additionally, subtype C viruses have been found to predominantly use CCR5 as a coreceptor, even in advanced AIDS stages, although CXCR4-tropic viruses can emerge with antiretroviral treatment (Johnston et al., 2003). HIV-1 subtype C appears to develop fewer drug resistance mutations compared to non-C subtypes, particularly subtype B. However, its global prevalence may be attributed to other factors such as transmission fitness rather than drug resistance patterns.

Our assay seamlessly detected HIV-1 along with its C subtype, as reflected by the FAM and SUN signals. Non-C subtypes were confirmed by the absence of any SUN signal despite the presence of a signal in the FAM channel. The use of GAPDH was primarily to assess PCR inhibition, given that the GAPDH target resides on human chromosome 12. Its detection was independent of the presence or absence of HIV-1 proviral DNA, making it an appropriate internal control for inhibition detection. This is particularly relevant since the analyte for HIV-1 proviral detection was human peripheral blood DNA, where the presence of human DNA serves as an intrinsic quality control.

The inclusion of a Stage 1 end-point PCR, which can also be carried out on the same real-time PCR platform, serves as an additional measure to enhance the sensitivity of the reaction. This step primarily enriches the diagnostic segment within the HIV-1 proviral stretch, providing the advantages associated with a nested PCR approach. The fluorescence-based qPCR in Stage 2 eliminates the need for laborious and time-consuming agarose gel electrophoresis while significantly improving the assay’s sensitivity through the use of dye-labeled probes.

A significant aspect of this assay is its reliance on a single-contact quenching technology, where quenching of the dye occurs in trans, as opposed to the classical TaqMan dual-labeled probes, where quenching occurs in cis. In the latter, the quencher is physically linked to the same oligonucleotide as the fluorophore, ensuring efficient energy transfer and quenching. In contrast, the FRET-based quenching technology used here relies on the proximity of the fluorophore and quencher, which are separated by approximately 17-18 bases, potentially affecting quenching efficiency.

A single homologous quencher-tagged oligonucleotide molecule facilitates quenching of all three dye-labeled probes, namely FAM, SUN (VIC), and Cy5, respectively. This design is cost-effective since the synthesis of a single-labeled probe is more economical compared to the synthesis of dual-labeled TaqMan probes.

Another important feature of this assay is the use of biotags— short 8 bp tags engineered at the 5’ end of the gene-specific primers. These tags allow fluorescent probes to recognize their target primers, ensuring accurate annealing and amplification to generate signals. The incorporation of biotags enhances the flexibility of multiplexing the assay while also improving the accuracy of target-specific signal interpretation.

The human GAPDH gene served as an internal control for PCR inhibition monitoring. Since the analyte in this assay is primarily human peripheral blood, where the occurrence of the GAPDH gene is inevitable, its detection is independent of the presence or absence of HIV-1 proviral DNA, ensuring reliable assay performance.

A small panel of 11 samples was subjected to a validation study for the protocol developed in this study. The samples were initially analyzed for HIV-1 drug resistance using a protocol developed by our group (Acharya et al., 2014). This analysis not only identified potential HIV-1 drug resistance mutations but also determined the virus subtype. Seven samples were confirmed as HIV-1 positive with subtype C, while three samples were HIV-1 positive with another subtype (B); one sample was from a healthy individual, bringing the total to 11 samples. When cross-tested using the protocol developed here, the fluorescence profiles of all samples matched those obtained by the gold standard HIV-1 nucleotide sequencing-based drug resistance test, except for one HIV-1 subtype C sample that could not be analyzed due to PCR inhibition, which prevented amplification of all targets in the reaction.

Multiplexing in HIV detection using qPCR has been demonstrated in several studies, allowing for simultaneous quantification of HIV proviral DNA, subtype identification, and inclusion of internal controls. The intact proviral DNA assay (IPDA) is a notable example of multiplexing in HIV detection. This assay uses multiplex digital droplet PCR to quantify intact proviruses while distinguishing them from defective ones (Simonetti et al., 2020). Interestingly, while multiplexing offers advantages in HIV detection, it can also present challenges. For instance, (Jaworski et al., 2018) highlights the variability in BLV proviral DNA copy number measurements among different laboratories using qPCR assays, emphasizing the need for standardization in multiplex assays. This observation underscores the importance of quality control and validation in multiplex HIV detection methods.

Multiplexing in HIV proviral DNA detection by qPCR enables comprehensive analysis, including quantification, subtype identification, and internal control validation. The IPDA exemplifies this approach by offering improved sensitivity and specificity in HIV reservoir quantification. However, to our knowledge, this is the first instance of developing a multiplex assay to detect HIV-1 along with its C subtype variant and with an internal control feature using dye-labeled probes quenched in trans, as opposed to the popular method of using dual-labeled probes where the dyes are quenched in cis.

## Supporting information

https://docs.google.com/document/d/1HB-Eyk26w9iNR81Eb32imHlSG6wWo36I/edit?usp=sharing&ouid=115148606976539336609&rtpof=true&sd=true

## Data Availability

All data produced in the present study are available upon reasonable request to the authors.

## Acknowledgement

The authors gratefully acknowledge Sn Genelab Private Limited for providing the HIV proviral DNA from clinical samples used in this study.

