## Supplementary material for "Multiplex qPCR Assay for HIV-1 Proviral DNA Detection and Subtype Characterization: Exploiting Quenching of Multiple Fluorophores with a Single Quencher Operating in *Trans*": https://docs.google.com/document/d/1HB-Eyk26w9iNR81Eb32imHlSG6wWo36I/edit?usp=sharing&ouid=115148606976539336609&rtpof=true&sd=true

Supplementary information

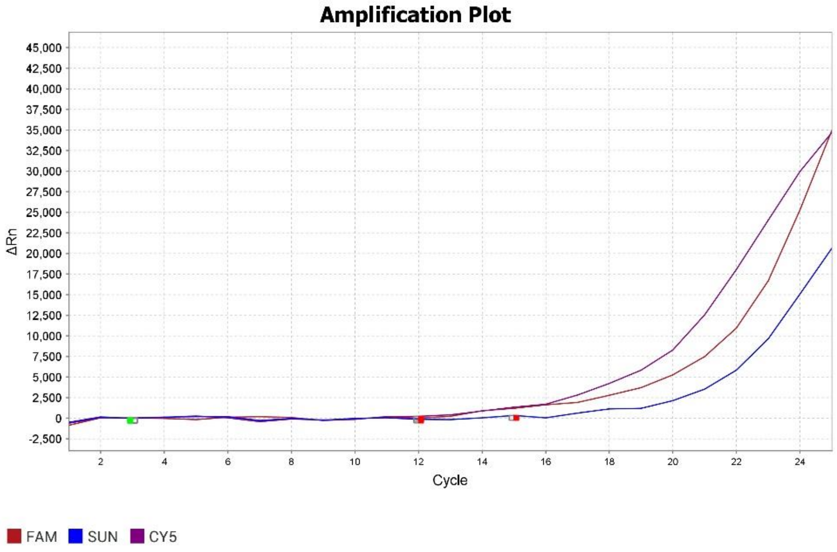


Sample No. 1 (HIV-1, C-Subtype)

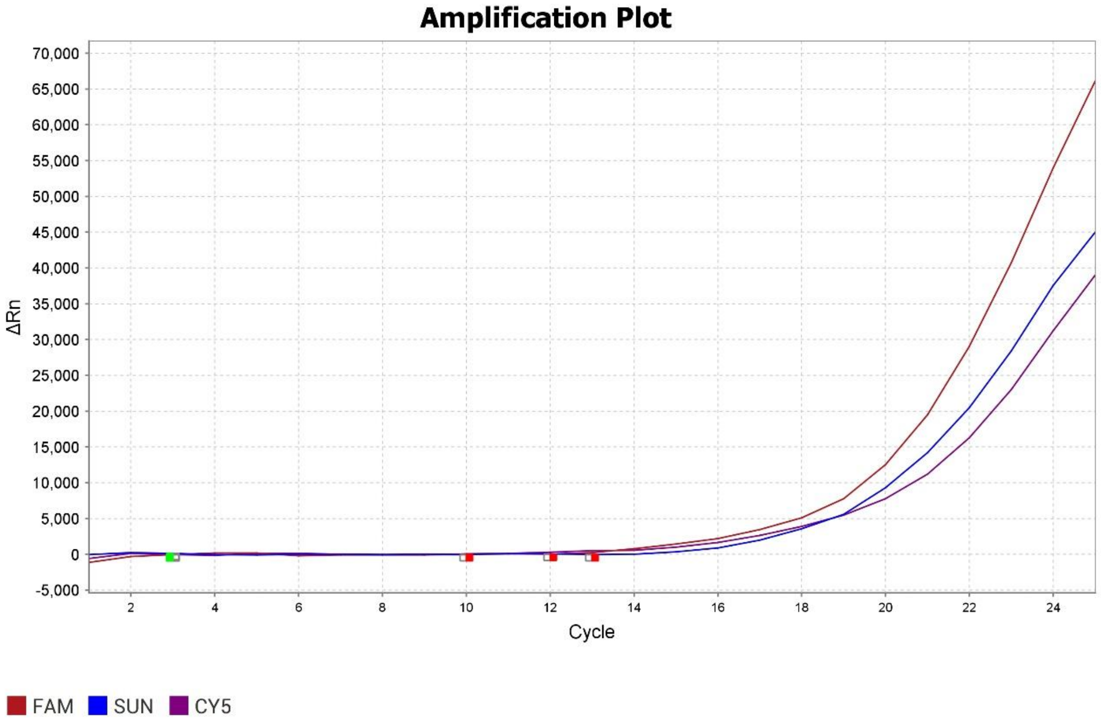


Sample No. 3 (HIV-1, C-Subtype)

Supplementary information

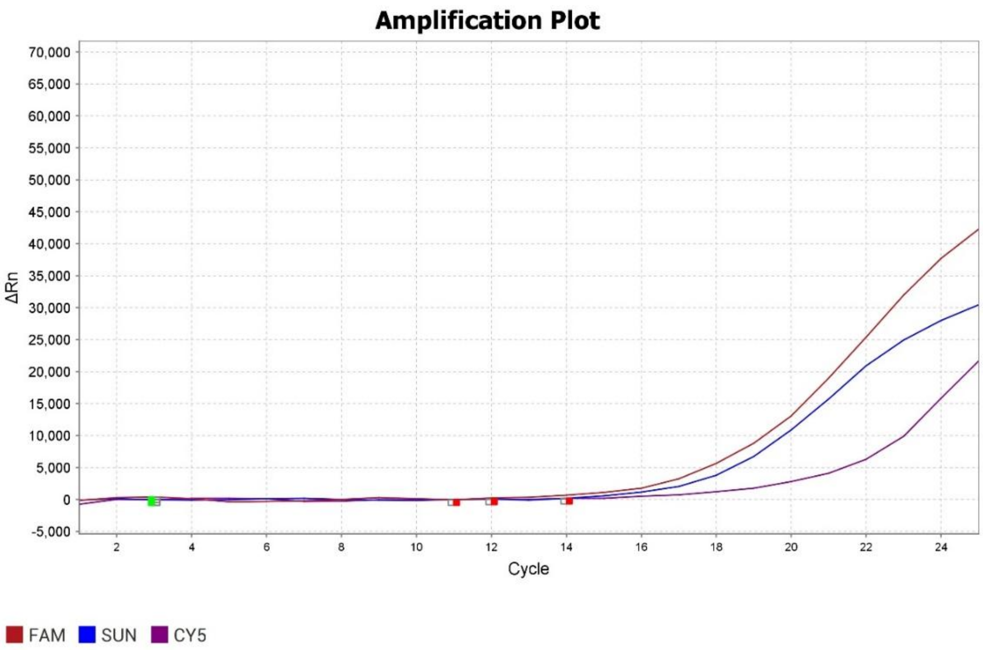


Sample No. 4 (HIV-1, C-Subtype)

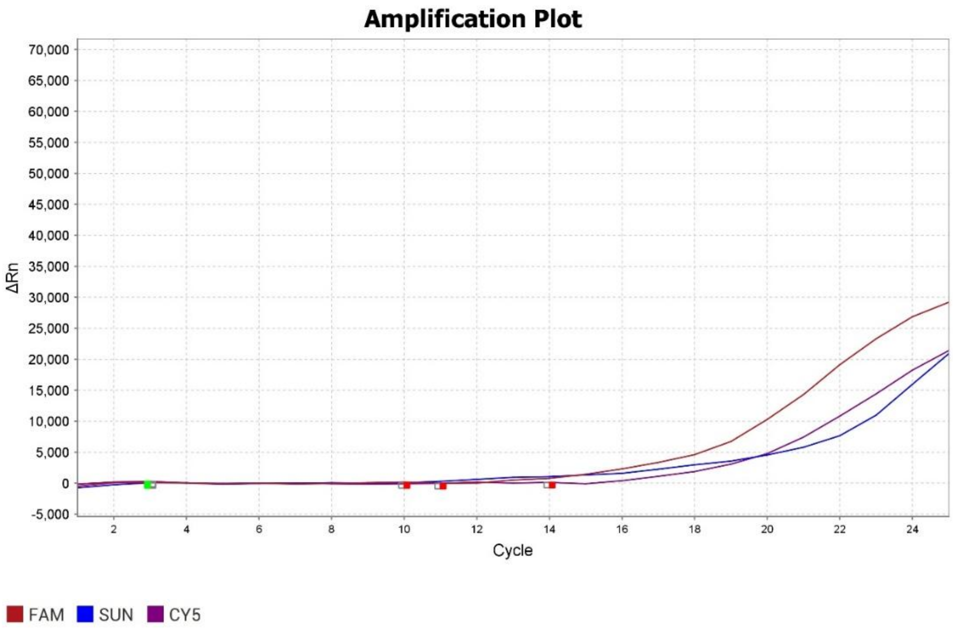


Sample No. 7 (HIV-1, C-Subtype)

Supplementary information

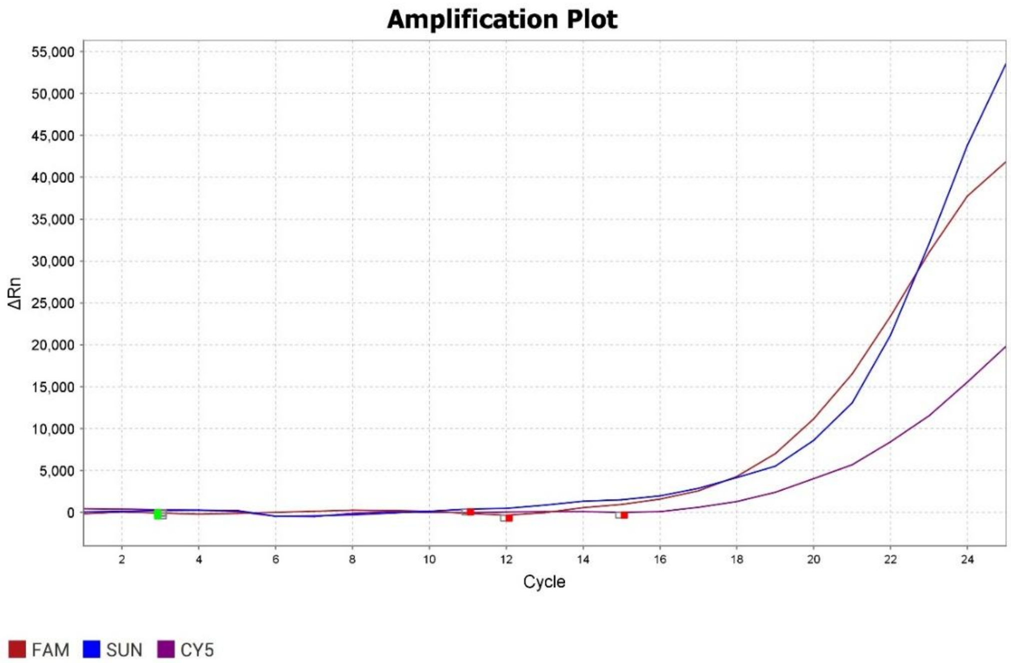


Sample No. 10 (HIV-1, C-Subtype)

Supplementary information

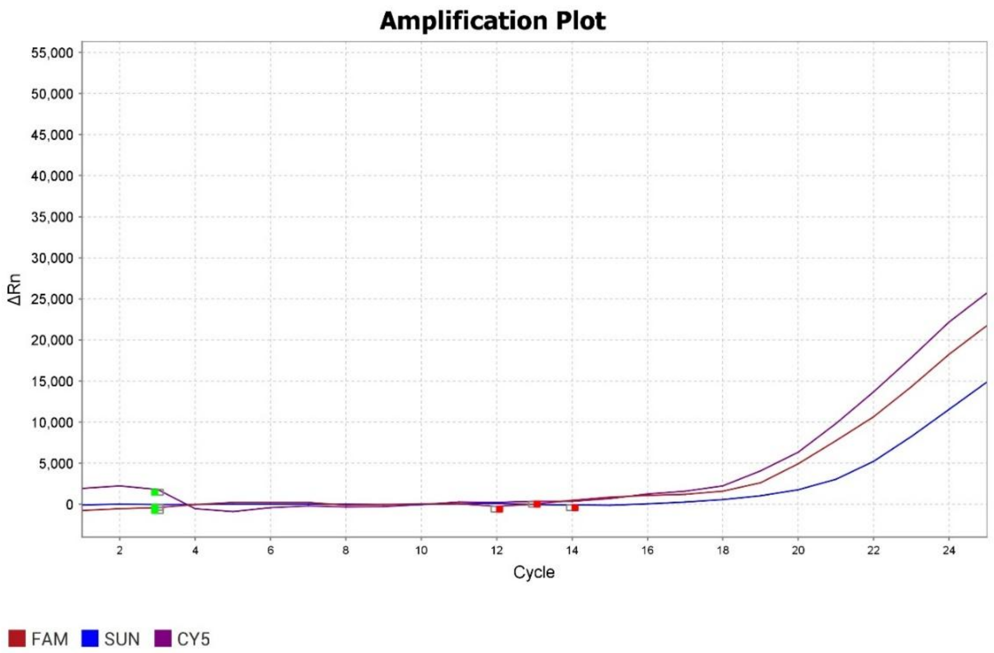


Sample No. 11 (HIV-1, C-Subtype)

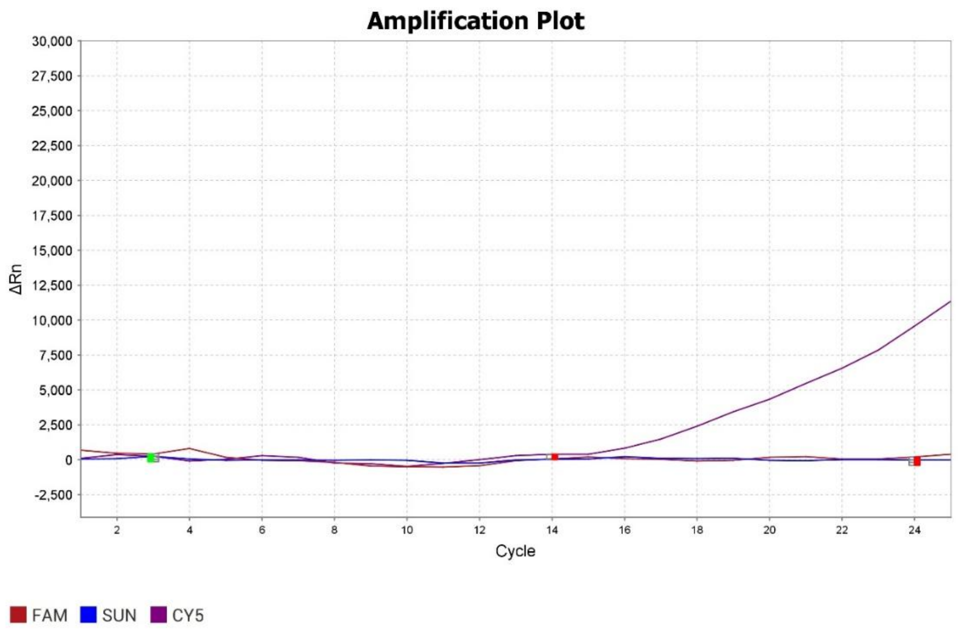


Sample No. 5 (Healthy Individual)

Supplementary information

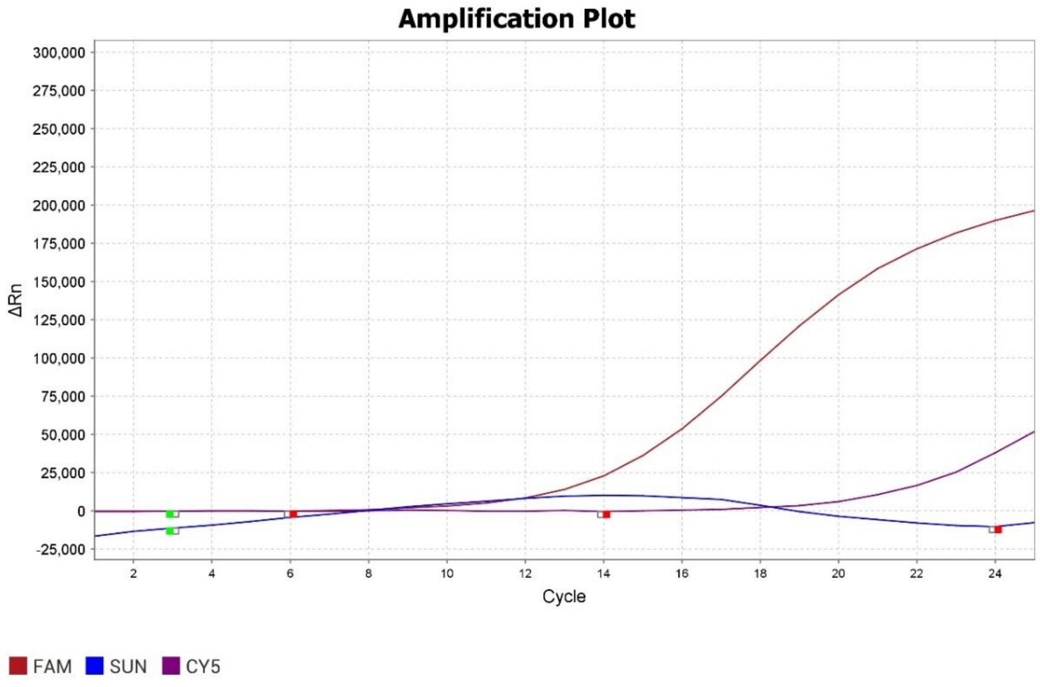


Sample No. 2 (HIV-1, B-Subtype)

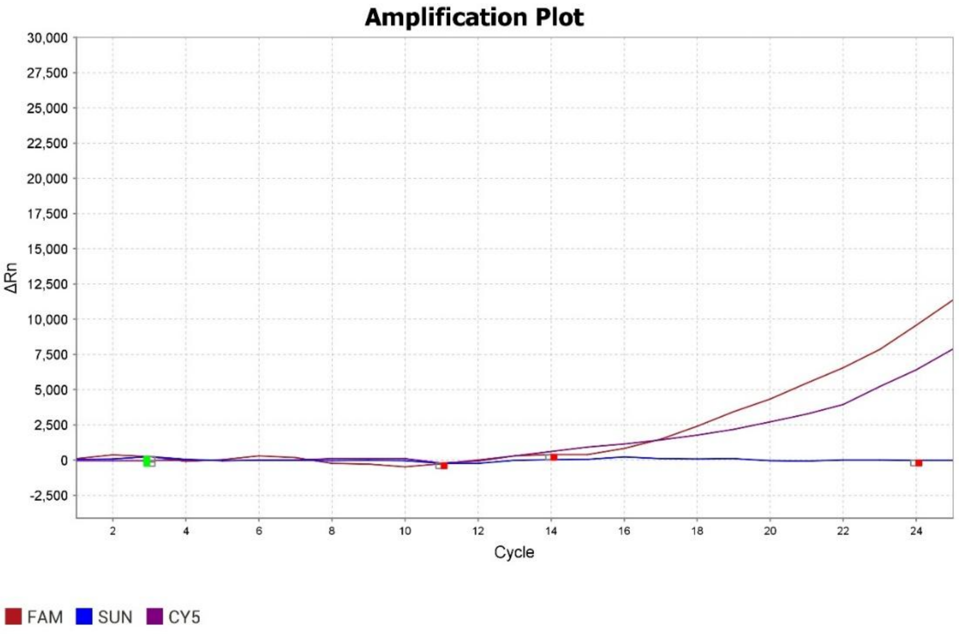


Sample No. 6 (HIV-1, B-Subtype)

Supplementary information

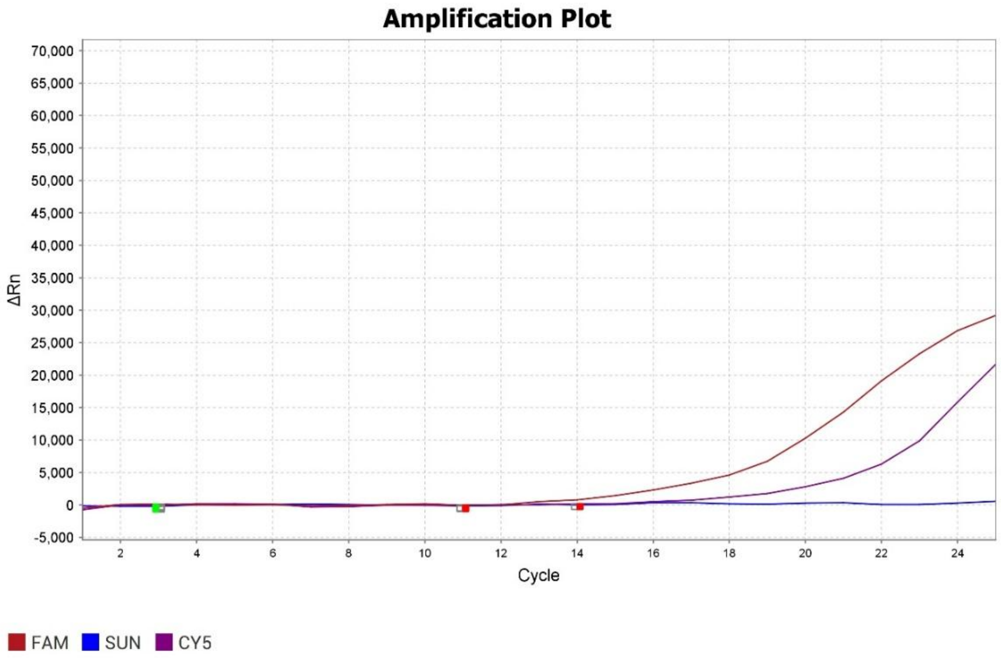


Sample No. 9 (HIV-1, B-Subtype)

Supplementary information

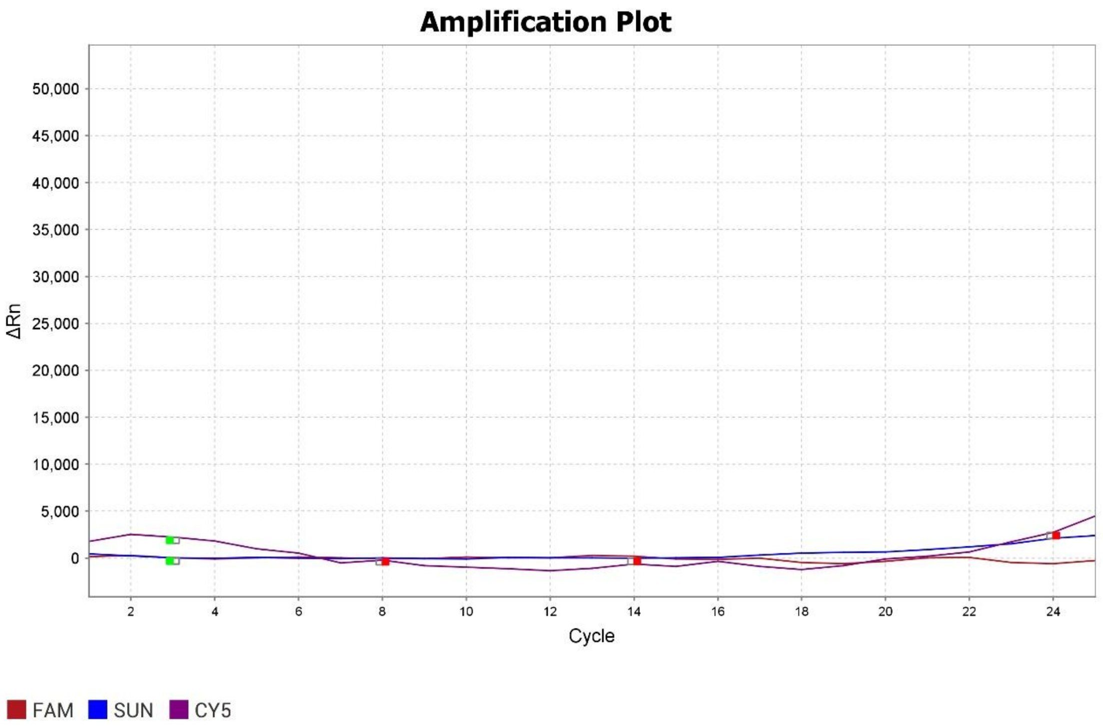


Sample No. 8 (PCR Inhibition)
